# Analytical performances of the COVISTIX™ and Panbio™ antigen rapid tests for SARS-CoV-2 detection in an unselected population (all commers)

**DOI:** 10.1101/2021.09.10.21263410

**Authors:** Francisco Garcia-Cardenas, Alba Franco, Ricardo Cortés, Jenny Bertin, Rafael Valdéz, Fernando Peñaloza, Emmanuel Frias-Jimenez, Alberto Cedro-Tanda, Alfredo Mendoza-Vargas, Juan Pablo Reyes-Grajeda, Alfredo Hidalgo-Miranda, Luis A. Herrera

## Abstract

**Importance:** A steady increase in acute respiratory syndrome coronavirus 2 (SARS-CoV-2) cases worldwide is causing some regions of the world to withstand a third or even fourth wave of contagion. Swift detection of SARS-CoV-2 infection is paramount for the containment of cases, prevention of sustained contagion; and most importantly, for the reduction of mortality.

**Objective:** To evaluate the performance and validity of the COVISTIX™ rapid antigen test, for the detection of SARS-CoV-2 in an unselected population and compare it to Panbio™ rapid antigen test and RT-PCR.

**Design:** This is comparative effectiveness study; samples were collected at two point-of-care facilities in Mexico City between May and August 2021.

**Participants:** Recruited individuals were probable COVID-19 cases, either symptomatic or asymptomatic persons that were at risk of infection due to close contact to SARS-CoV-2 positive cases.

**Diagnostic intervention:** RT-PCR was used as gold standard for detection of SARS-CoV-2 in nasal and nasopharyngeal swabs, study subjects were tested in parallel either with the COVISTIX™ or with Panbio™ rapid antigen test.

**Main outcome:** Diagnostic performance of the COVISTIX™ assay is adequate in all commers since its accuracy parameters were not affected in samples collected after 7 days of symptom onset, and it detected almost 65% of samples with a Ct-value between 30 and 34.

**Results:** For the population tested with COVISTIX™ (n=783), specificity and sensitivity of the was 96.0% (CI95% 94.0-98.0) and 81% (CI95% 76.0-85.0), as for the Panbio™ (n=2202) population, was 99.0% (CI95%: 0.99-1.00) and 62% (CI%: 58.0-64.0%), respectively.

**Conclusions and relevance:** The COVISTIX™ rapid antigen test shows a high performance in all comers, thus, this test is also adequate for testing patients who have passed the peak of viral shedding or for asymptomatic patients.

## Introduction

The Coronavirus Disease 2019 (COVID-19) pandemic has exerted unprecedented effects on healthcare and economic systems globally. A steady increase in acute respiratory syndrome coronavirus 2 (SARS-CoV-2) cases worldwide is causing some regions of the world to withstand a third or even fourth wave of contagion. (1,2) Swift detection of SARS-CoV-2 infection is paramount for the containment of cases, prevention of sustained contagion to return to economic and education activities; and most importantly, for the reduction of mortality. (3,4)

Reverse transcription polymerase chain reaction (RT-PCR) is considered the gold standard method for detecting SARS-CoV-2 infection due to its high sensitivity and specificity (5,6). However, implementing RT-PCR testing in massive screening campaigns requires specialized protective equipment, qualified personnel, and sample transportation to a centralized laboratory, which has proven to be challenging, particularly in resource-limited settings (2,7). As a result of these limitations, several rapid tests based on SARS-CoV-2 antigen detection by immunochromatography have been introduced, offering improved access to testing due to faster result availability, simple use at point-of-care, and low costs. Rapid antigen tests represent an appealing alternative for large-scale testing of the general population (8,9).

As an essential part of the public health response to the COVID-19 pandemic, Mexico City’s government implemented a surveillance strategy intended to detect active cases among the general population, which was initially based on RT-PCR. In November 2020, rapid antigen tests were also included in the strategy. According to data reported by Mexico City’s Digital Agency of Public Innovation (ADIP), almost 1 million antigen tests have been performed by the beginning of February 2021(10).

Although rapid technological advances in automated portable assays have allowed testing to be performed outside laboratory ambiences, these technologies have not shown a similar sensitivity compared to the efficient but highly consuming molecular assays performed in laboratory settings, increasing the risk of false negative results (11,12). Two inherent characteristics should be considered when considering which test to use; sensitivity and specificity which provide information on the accuracy of the test to measure the absence or presence of the disease. Nonetheless, the adequacy of a test in a certain population is elucidated by the positive predictive value and negative predictive value. These last measures vary depending on the true prevalence of the disease in a population as they evaluate the probability that a person with a positive or negative test result truly has or does not have the disease respectively. Thus, parameters such as the likelihood ratio, which is not influenced by the prevalence, should be taken into consideration in this pandemic since prevalence fluctuates across populations and over time (9).

The World Health Organization (WHO) recognizes that although antigen tests have proven lower sensitivity than molecular tests, they provide rapid and less resource-consuming means of detection of SARS-CoV-2 in individuals who have high viral loads; therefore, have higher risk of disease transmission (13). Currently, Mexico’
ss Institute of Epidemiologic Reference and Diagnosis (InDRE) has evaluated and approved more than 20 rapid antigen tests used for SARS-CoV-2 detection, including Abbott’s Panbio**™** and Sorrento’s COVISTIX™ COVID-19 Ag Rapid Test Device (14). These single use devices are immunochromatographic assays that detects the SARS-COV-2 nucleocapsid protein providing results of active infection within 20 minutes; being quicker than RT-PCR procedure (15).

Panbio**™** COVID-19 Ag rapid tes t device has been proved and compared to other rapid antigen tests in order to assess its diagnostic performance in symptomatic and asymptomatic populations. The sensitivity of this test has been directly correlated to the magnitude of viral load in nasopharyngeal region, which might exhibit inconsistencies in these populations due to the behavior of SARS-CoV-2 load in the upper respiratory tract owing to the viral load peak timing is uncertain in asymptomatic patients (16,17). Another limitation of this test is that it shows major sensitivity within the first 5-7 days after symptom onset depending on the viral load in the specimen used for evaluation; notwithstanding those patients with COVID-19 can remain positive for 2-3 weeks after the commencement of symptoms (11,18). Hence, the return to work of essential workers or traveling permissions might be misconceived.

Different strategies for mitigating contagion using antigen rapid device test have been proposed such as the use only in symptomatic patients within 5-7 days after symptom onset due that the viral load is at its peak. Furthermore, strategies such as the implementation of these assays to detect SARS-CoV-2 in healthcare workers and contacts of confirmed cases could be beneficial for pandemic containment (9). Therefore, the need for an economic yet precise testing strategy and a more sensitive antigen test that can rapidly detect lower viral loads at the very beginning of the disease or after 8 days after the start of the symptoms has emerged.

Here, we set to evaluate the performance and validity of the Sorrento’s COVISTIX™ rapid antigen test, for the detection of SARS-CoV-2 in nasal and nasopharyngeal swabs in a Mexican open population and compare it to Panbio™ rapid antigen test and RT-PCR.

## Methods

This study was conducted by the National Institute of Genomic Medicine of Mexico (INMEGEN), in collaboration with the Citibanamex COVID temporal unit set in Mexico City. Samples from seven-hundred eighty-three individuals were collected at the Citibanamex COVID temporal unit and at INMEGEN, Mexico City between May 1^st^ and August 16^th^, 2021; nasal and nasopharyngeal swabs were obtained and tested with the COVISTIX™ rapid antigen test according to manufacturer’s indications, as well as with RT-PCR. The algorithm implemented for testing with the COVISTIX™ rapid antigen test was to assess first a nasal swab, if negative, a second test was performed with a nasopharyngeal swab sample, if negative, the subject was considered as negative for SARS-CoV-2. Subjects were considered as positive for SARS-CoV-2 if either the nasal or the nasopharyngeal swab resulted positive with the COVISTIX™ rapid antigen test. In all individuals, results were confirmed by RT-PCR in a nasopharyngeal sample taken in parallel.

Additionally, we collected nasopharyngeal swabs from 2202 individuals at INMEGEN, and were tested for SARS-CoV-2 with the Panbio™ COVID-19 rapid antigen test according to manufacturer’s instructions and compared to RT-PCR in a nasopharyngeal sample taken in parallel. The study was approved by the ethics and research committees of Instituto Nacional de Medicina Genómica (CEI/1479/20 and CEI 2020/21).

### RNA extraction and RT-PCR detection

Total nucleic acids were extracted from 300 μL of viral transport media from the nasopharyngeal swab using the MagMAX Viral/Pathogen Nucleic Acid Isolation Kit (ThermoFisher Scientific) following the manufacturer’s instructions and eluted into 75 μL of elution buffer. RT-PCR was performed using the TaqPath™ COVID-19 CE-IVD RT-PCR Kit (ThermoFisher Scientific) following manufacturer’
ss instructions. Briefly, the kit detects the ORF1ab, S and N genes of the virus. We classified samples as positive for SARS-CoV-2 when primer-probe sets were detected with a Ct-value of less than 40. If only one of these genes was detected, we labeled the sample as inconclusive. We ran all tests with Thermo Fisher’s ABI QuantStudio 5 or QuantStudio 7 real-time thermal cyclers.

### Statistical Analysis

A 2×2 table was built using RT-PCR as the gold standard. Sensitivity, specificity, likelihood ratios, post-test probabilities, Cohen’s *kappa* correlation coefficient and Youden index were calculated for both rapid antigen assays.

## Results and Discussion

A total of 783 subjects were included to evaluate the COVISTIX™ assay, 254 of which had a positive RT-PCR test (prevalence 32.4%). In this group of individuals, 391 were female and 392 were male. The median (IQR) age in years was 40 (28-51). Nasal and nasopharyngeal samples were evaluated with the COVISTIX™ rapid antigen test and compared to nasopharyngeal swab analyzed with RT-PCR. Table 1 shows that out of 783 samples, 205 tested positive both by RT-PCR and by COVISTIX™ and 508 were detected negative by both assays, showing false negative results in 49 samples (19.3%) and 21 false positive results (4%). Overall specificity and sensitivity of the COVISTIX™ rapid antigen test was 96% (CI95%: 94-98) and 81% (CI95%: 76-85), respectively. Positive and negative likelihood ratios were 20.25 (CI95%: 13.0-31.0) and 0.2 (CI95%: 0.16-0.26) each. Positive post-test probability was 91% and negative post-test probability was 12%. Cohen’s Kappa coefficient shows a very good concordance between results obtained by COVISTIX™ and RT-PCR (0.8; CI95%: 0.72-0.86).

**Table 1.** Results of **COVI-STIX™** SARS-CoV-2 antigen rapid test in swabs from **783 individuals** (all commers) compared to RT-PCR.

|  |  |  |  |
| --- | --- | --- | --- |
| Sex | Female: 391 (49.9%) |  | Male: 392 (50.1%) |
| Median age | 40 years (IQR: 28-51) |  |  |
|  | PCR (+) | PCR (-) | Total |
| COVI-STIX (+) | 205 | 21 | 226 |
| COVI-STIX (-) | 49 | 508 | 557 |
| Total | 254 | 529 | 783 |
| Prevalence | 0.32 |  |  |
| Sensitivity | 0.81 (CI95%: 0.76-0.85) |  |  |
| Specificity | 0.96 (CI95%: 0.94-0.98) |  |  |
| LR (+) | 20.25 (CI95%: 13-31) |  |  |
| LR (-) | 0.2 (CI95%: 0.16-0.26) |  |  |
| Post-Test (+) | 0.91 (CI95%: 0.86-0.94) |  |  |
| Post-Test (-) | 0.12 (CI95%: 0.07-0.11) |  |  |
| Kappa | 0.8 (CI95%: 0.72-0.86) |  |  |
| Youden index | 0.77 (SE: 0.026) |  |  |
| LR: Likelihood Ratio; Post-Test: Post-Test Probability.<br>Kappa: Cohen's Kappa correlation (Very good) |  |  |  |

One of the most frequently used antigen rapid tests worldwide during pandemic is the Panbio™ rapid antigen test. In this study we compared the performance of the COVISTIX™ assay with Panbio™’s. A total of 2202 subjects were included, 443 of which had a positive RT-PCR test (prevalence 20%). In this cohort, 979 patients were female and 1223 were male. The median (IQR) age in years was 41 (30-50) (Table 2). Out of 2202 patients, 275 tested positive both by RT-PCR and by Panbio™ and 1755 performed negative for both assays. Discordant results (false negatives) are shown in 168 (37.9%) and 4 (0.23%) false positive results with Panbio™; overall specificity and sensitivity was 99% (CI95%: 99.0-1.0) and 62% (CI%: 58.0-64.0%), respectively. Positive and negative likelihood ratio were 273 (CI95%: 102-279) and 0.38 (CI95%: 0.34-0.43) each. Positive post-test probability was 99% (CI95%: 96–99) and for negative post-test probability was 9% (CI95%: 8-10). Cohen’
ss Kappa test was calculated showing a correlation of 0.7 (CI95%: 0.7-0.8).

**Table 2.** Results of **Panbio™** SARS-CoV-2 antigen rapid test in swabs from **2202 individuals** (all comers) compared to RT-PCR.

|  |  |  |  |
| --- | --- | --- | --- |
| Sex | Female: 979 |  | Male: 1223 |
| Median age | 41 years (IQR: 30-50) |  |  |
|  | PCR (+) | PCR (-) | Total |
| Panbio (+) | 275 | 4 | 279 |
| Panbio (-) | 168 | 1755 | 1923 |
| Total | 443 | 1759 | 2202 |
| Prevalence | 0.20 |  |  |
| Sensitivity | 0.62 (CI95%: 0.58-0.64) |  |  |
| Specificity | 0.99 (CI95%: 0.99-1.00) |  |  |
| LR (+) | 273 (CI95%: 102-279) |  |  |
| LR (-) | 0.38 (CI95%: 0.34-0.43) |  |  |
| Post-Test (+) | 0.99 (CI95%: 0.96-0.99) |  |  |
| Post-Test (-) | 0.09 (CI95%: 0.08-0.10) |  |  |
| Kappa | 0.72 (CI95%: 0.68-0.76) |  |  |
| Youden index | 0.62 (SE: 0.023) |  |  |
| LR: Likelihood Ratio; Post-Test: Post-Test Probability.<br>Kappa: Cohen's Kappa correlation (Good) |  |  |  |

In order to evaluate the overall accuracy of the COVISTIX™ assay for detection of SARS-CoV-2 carriers, we calculated the Youden index (J), which is a useful measure of the misclassification error in diagnostic tests (19). The J value for COVISTIX™ was 0.77 (CI95%: 0.72-0.82; SE: 0.026) indicating a good performance of this assay. Additionally, we compared this result with the observed with the Panbio™ test, J: 0.62 (CI95%: 0.57-0.67; SE: 0.023). Since the CI95% do not overlap, we can conclude that the difference observed between the indexes is real, and significant with a *t-*test value of 4.3.

Several reports have emphasized the importance of the viral load for the detection of SARS-CoV-2 carriers by rapid antigen tests, which are best-suited for the rapid identification of individuals carrying high viral loads(12,20). RT-PCR Ct-value is considered a surrogate parameter for viral load, the lower the Ct-value the higher the expected viral load, therefore we evaluated the performance of the COVISTIX™ rapid antigen test based on the RT-PCR Ct-value. Although assay’s sensitivity was higher in samples with a Ct-value below 30, as it has been reported for other rapid antigen tests, the COVISTIX™ assay detected almost 65% of SARS-CoV-2 carriers with Ct-values between 30 and 34 (Table 3A), which was not observed with the Panbio™ rapid antigen test (Table 3B). These results indicate that the COVISTIX™ assay is effective to detect not only highly infectious individuals but also those potentially carrying low viral loads.

**Table 3.**
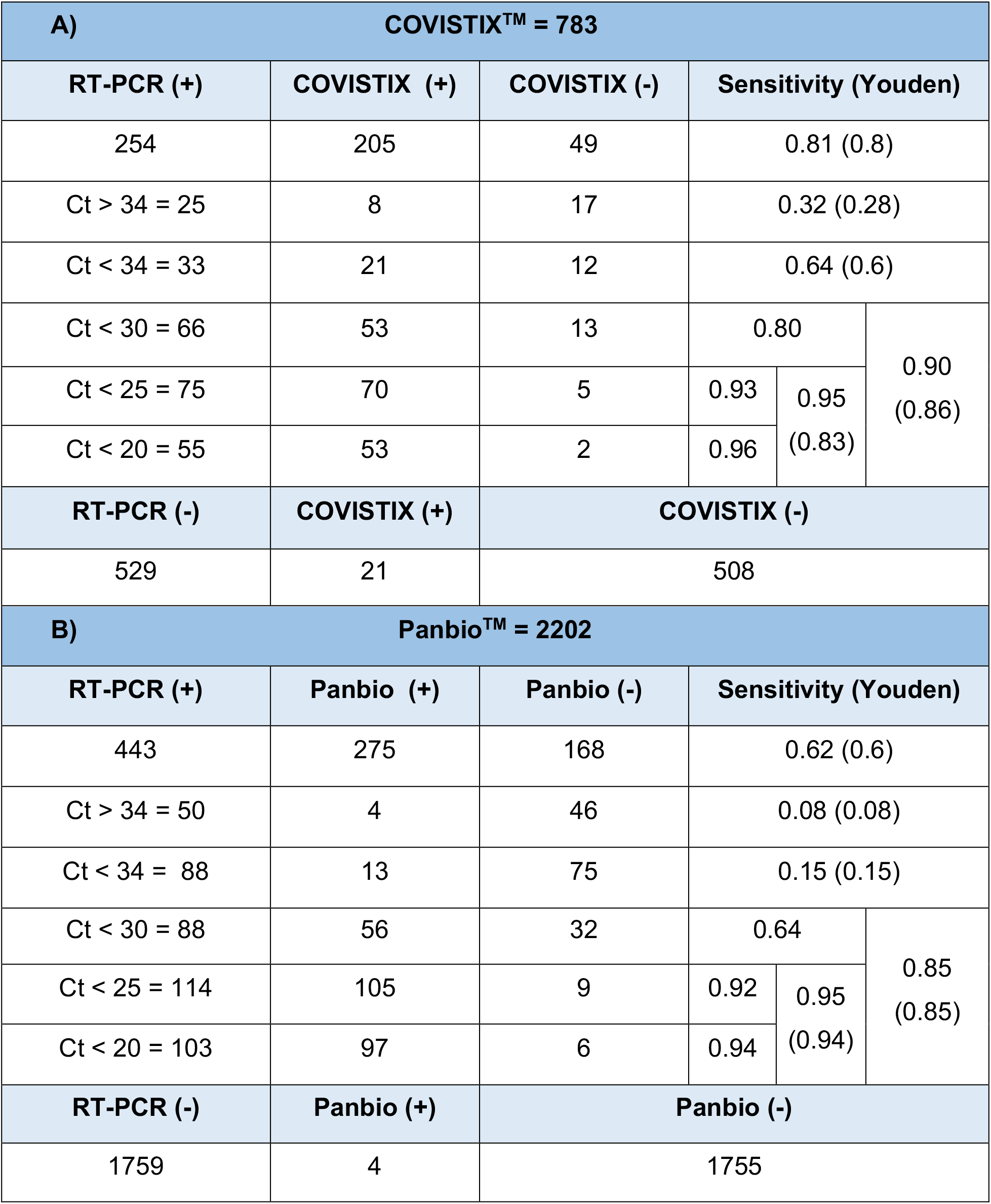
COVISTIX™ and Panbio™ results compared to RT-PCR (N = 783 and 2202 respectively) depending on the Ct-value.

We also analyzed the performance of the COVISTIX™ assay in symptomatic (N=335) and asymptomatic (N=448) individuals. Results show an overall sensitivity very similar among symptomatic individuals regardless of the number of days after symptom onset (Table 4A). The sensitivity observed in the asymptomatic group was lower than in the symptomatic group (Table 4B), however, the difference was not significant when we compared the J indexes of both groups (*t*-test: 0.30).

**Table 4.** Overall performance of the COVISTIX™ rapid antigen test in symptomatic and asymptomatic individuals.

| A) Symptomatic = 335 |  |  |  |
| --- | --- | --- | --- |
| ≤ 7 DAYS | RT-PCR (+) | RT-PCR (-) | Sensitivity (CI 95%)<br>(Youden index) |
| COVISTIX (+) | 103 | 12 | 0.82 (0.76-0.89)<br>(0.72) |
| COVISTIX (-) | 22 | 103 |  |
| > 7 DAYS | RT-PCR (+) | RT-PCR (-) | Sensitivity (CI 95%)<br>(Youden index) |
| COVISTIX (+) | 56 | 4 | 0.82 (0.73-0.91)<br>(0.67) |
| COVISTIX (-) | 12 | 23 |  |
| B) Asymptomatic = 448 |  |  |  |
|  | RT-PCR (+) | RT-PCR (-) | Sensitivity (CI 95%)<br>(Youden index) |
| COVISTIX (+) | 46 | 5 | 0.75 (0.65-0.86)<br>(0.74) |
| COVISTIX (-) | 15 | 382 |  |

A major concern of the use of rapid antigen tests for massive screening or even for diagnosis is the elevated frequency of false negatives, whose impact in pandemic control could be detrimental since false-negative individuals could spread the virus due to an unjustified sense of security (21). In general, several studies suggest that rapid antigen tests are frequently negative in RT-PCR positive samples with Ct-values above 29, which could lead to an elevated number of undetected SARS-CoV-2 carriers; since there is no minimal infectious dose reported to date it cannot be assumed that individuals whose samples report Ct-values above 30 are not contagious (22,23). In fact, La Scola et al reported that 50% of clinical specimens with Ct-value equal or more than 30 can be cultured and be potentially infectious (24). The present results show that performance of the COVISTIX™ rapid antigen test is adequate even if sampling is not restricted to individuals within the first seven days period of symptom onset nor to low viral load. Thus, this test could be a powerful tool for surveillance purposes in settings were RT-PCR is not easy to implement.

## Data Availability

The data is available within the manuscript.

